# Dental health of childhood cancer survivors – a report from the Swiss Childhood Cancer Survivor Study (SCCSS)

**DOI:** 10.1101/2024.09.24.24314228

**Authors:** C Nigg, C Matti, P Jörger, AO von Bueren, C Filippi, T Diesch-Furlanetto, Z Tomášiková, CE Kuehni, G Sommer

**Affiliations:** Institute of Social and Preventive Medicine, University of Bern, Switzerland; Graduate School for Health Sciences, University of Bern, Switzerland; Department of Paediatrics, Obstetrics and Gynaecology, Division of Paediatric Haematology and Oncology, University Hospital of Geneva, Switzerland; Department of Children and Adolescent Dentistry, University Center for Dental Medicine Basel (UZB), Basel, Switzerland; Department of Oncology/Haematology, Basel, University Children’s Hospital Basel UKBB, Basel, Switzerland; Childhood Cancer Switzerland; Division of Pediatric Hematology/Oncology, Inselspital, Bern University Hospital, University of Bern, Bern, Switzerland

**Keywords:** pediatric oncology, survivorship, late effects, dental care, cohort study, late effects

## Abstract

**Background:** Cancer and its treatments can affect dental health of childhood cancer survivors. We aimed to evaluate prevalence of dental problems in survivors, compare them to their siblings, and investigate cancer-related risk factors.

**Methods:** As part of the population-based Swiss Childhood Cancer Survivors Study, we sent questionnaires inquiring about dental problems to survivors aged 5-19 years and their siblings. We retrieved cancer-relevant information from the Swiss Childhood Cancer Registry and used logistic regressions to compare dental problems between survivors and siblings and to investigate cancer-related risk factors.

**Results:** We included 735 survivors and 144 siblings. Almost half of survivors and siblings reported at least one dental problem. Compared to siblings, survivors had a greater risk for hypo- or microdontia (OR 1.7; 95%CI 0.9-3.2) and enamel hypoplasia (2.2; 0.8-6.0), but similar risk for cavities or cavity-related tooth loss (0.8; 0.6-1.3). Chemotherapy was associated with enamel hypoplasia (3.0; 1.2-10.4), cavities or cavity-related tooth loss (1.5; 1.0-2.3), and gum problems during (23.0; 9.4-76.2) and after (4.6; 2.0-13.5) treatment. Hematopoietic stem cell transplantation (HSCT) was related to hypo- or microdontia (5.4; 2.6-10.7), cavities or cavity-related tooth loss (2.1; 1.2-3.6), and gum problems during treatment (2.0; 1.2-3.6). For hypo- or microdontia and cavities, associations with treatment were driven by patients diagnosed <5 years.

**Conclusion:** Childhood cancer patients treated with chemotherapy or HSCT, especially at a young age, were at higher risk for dental problems. Regular dental check-ups guided by healthcare teams and dental hygiene habits can mitigate risks and promote survivor’s long-term dental health.

## Introduction

Medical advancements allow to cure most children diagnosed with cancer, with 5-year survival rates approaching 90% in Switzerland.^1^ Despite those advancements, two thirds of survivors develop somatic late effects of cancer treatments that may include a broad range of health problems.^2–7^ Some late effects are already visible shortly after treatment, including dental problems.^8^ Early childhood is a crucial phase for crown and root formation,^9^ making patients treated with radiation to the head, chemotherapy, or hematopoietic stem cell transplantation (HSCT) particularly susceptible for dental late effects. Radiation to the head and neck area can cause salivary gland hypofunction, resulting in xerostomia and increased caries,^10,11^ or damage the tooth bud, disrupting tooth development and leading to structural anomalies, such as enamel hypoplasia, microdontia, or hypodontia.^12^ Chemotherapy, especially alkylating agents, interfere with the cell cycle and the intracellular metabolism, altering odontoblast and ameloblast activity.^11,13^ Recipients of HSCT may experience salivary gland hypofunction as a result of conditioning regimes or as an early symptom of chronic graft-versus-host disease.^11,14,15^

Two previous meta-analyses including children treated with chemotherapy or a combination of chemotherapy and radiotherapy indicate an increased risk for tooth agenesis and microdontia, enamel hypoplasia, cavities, and gingivitis.^8,16^ While most of the studies included used dental examinations, they were restricted to patients receiving specific treatment regimes, or small populations. The few population-based or cohort studies on dental health come from North America and only include adult participants^12,17^, who were already in their thirties or forties at time of study. We aimed to evaluate dental health in a population-based study of young childhood cancer survivors (CCS) in Switzerland, a country with different dental hygiene practices and eating habits than North America.^18,19^ In particular, this study evaluated prevalence of dental problems in pediatric CCS, compared it with their siblings, and investigated cancer-related factors contributing to dental problems in pediatric CCS.

## Methods

### Study design and study population

The Swiss Childhood Cancer Survivor Study (SCCSS) is a population-based study nested in the Swiss Childhood Cancer Registry (ChCR). Detailed methods of the SCCSS are available elsewhere.^20^ The ChCR includes all children and adolescents diagnosed with leukemias, lymphomas, central nervous system (CNS) tumors, malignant solid tumors, or Langerhans cell histiocytosis prior to age of 20 years in Switzerland since 1976.^21^ As part of the SCCSS, we sent questionnaires to CCS who had survived ≥5 years since their initial diagnosis. Questionnaires are available in German, French, and Italian language. For this study, we included CCS who participated in the SCCSS between 2015 and 2022, were aged between 5 and 19 years at the time of study, and consented to participate. For CCS aged between 5 and 15 years, we asked their parents to fill-in the questionnaire. Siblings of similar age (5-19 years) received the same questionnaire between 2022 and 2024 without cancer-related questions. Ethical approval was granted by the ethics committee of the canton of Bern, Switzerland (KEK-BE: 166/2014 and 2021-01462).

### Dental health

The questionnaire asked CCS and siblings about experiencing hypodontia, microdontia, enamel hypoplasia, cavities, and tooth loss due to cavities and gum disease. It asked CCS, but not siblings, if they experienced gum problems during or after cancer treatment, and if they had problems with health insurance coverage for any dental problems. Participants could report on other dental problems qualitatively in a comment field. For a detailed overview of the questions, please see *Supplementary Table 1*. For questions in the original language, please contact the authors.

### Explanatory variables

#### Sociodemographic characteristics

We assessed the following sociodemographic characteristics via questionnaires: Swiss language region, sibling status, parental and participant nationality, and parental education. We categorized the language region into German and French/Italian. We divided CCS’ sibling status into yes (≥1 sibling) and no (0 siblings). We used the nationality to define migration background. For migration background and parental education, we applied definitions of the Federal Statistics Office.^22,23^ If both parents were born Swiss or if the participant and at least one parent were born Swiss, we defined this as no migration background.^22^ We considered the highest educational degree obtained by either parent, which we divided into three categories: primary education (compulsory schooling only), secondary education (higher schooling and vocational training), or tertiary education (upper vocational education, university or technical college).^23^ We received date of birth and sex of the CCS from the ChCR and of the siblings from questionnaires.

#### Cancer-related characteristics

We received the following cancer-related characteristics from the ChCR: age at diagnosis, diagnosis according to the International Classification of Childhood Cancer, third edition (ICCC-3)^24^ and Langerhans Cell Histiocytosis. Treatment information included chemotherapy, radiotherapy, cranial radiation, including radiation to the head, neck or total body irradiation with any dose, HSCT, and relapse (all yes/no).

### Statistical analysis

We first weighted siblings using inverse probability weighting to be representative of CCS with regards to sex, age, migration, and language region as described previously.^6,25^ Secondly, we calculated number and proportion of CCS and their siblings who reported at least one dental problem, as well as for each specific dental problem individually. Third, we used multivariable logistic regression analysis to compare the odds for dental problems between CCS and siblings. For this analysis, we combined hypo- and microdontia into one outcome and cavities and cavity-related tooth loss a second one. We considered adjusting for family clustering via multilevel modeling. Since one-level models yielded similar results, we decided for one-level models due to model parsimony. We computed separate multivariable logistic regression models for each dental problem, deciding *a priori* to include participant type (CCS/sibling), age at study, and sex as predictors. Fourth, to investigate cancer-related risk factors for each dental problem, we applied multivariable logistic regression analysis using data from CCS. We decided *a priori* to include chemotherapy, cranial radiation, HSCT, age at diagnosis, age at study, time since diagnosis, and relapse in all models, but dropped age at study due to multicollinearity. Since early childhood is a crucial dentition phase, we ran age-stratified analysis (diagnosis <5 years / ≥5 years) for prevalence of dental problems and investigation of cancer-related risk factors.^11^

We included all participants responding to at least one dental health question (complete-case analysis). In sensitivity analyses, we imputed missing values using the Multivariate Imputation via Chained Equations (MICE) package ^26^ to specify an imputation model with the algorithm iteratively imputing multiple possible values for missing values. We ran imputations separately for models comparing CCS and siblings, and for models investigating cancer-related risk factors in CCS. We generated 20 datasets with 20 iterations and pooled results based upon Rubin’s rule.^27^

## Results

In total, of 1597 families with eligible CCS, 1339 were contacted with a questionnaire. Of those, 763 (57%) returned the questionnaire and 735 (55%) responded to at least one question about dental problems. Of 451 families with eligible siblings, 438 were contacted. Of those, 144 responded to at least one question about dental problems (33%). *Supplementary Figure 1* shows the study population flow chart with details on reasons for not contacting or exclusion of CCS and siblings. Median age at study was 12.8 years (interquartile range [IQR] 10.2-15.2) for CCS and 13.5 years (IQR 11.1-15.5) for siblings. For CCS, median age at diagnosis was 4.2 years (IQR 2.1-7.2), and median time since diagnosis 8.2 years (IQR 6.9-9.4). Leukemia was the most common cancer diagnosis (41%), followed by CNS tumors (17%) and lymphomas (8%). Eighty percent of CCS had received chemotherapy treatment, 9% cranial radiotherapy, and 9% HSCT (see *Table 1*).

**TABLE 1.** Characteristics of childhood cancer survivors and siblings.

| Socio-demographic characteristics | Survivors<br>(N = 735) |  | Siblings<br>(N = 144) |  |
| --- | --- | --- | --- | --- |
|  | N | % | N | %<sup>a</sup> |
| Age at study (mean, standard error) | 13.2 (0.1) |  | 13.5 (0.3) |  |
| (median, IQR) | 12.8 [10.2– 15.4] |  | 13.3 [11.1–15.5] |  |
| Age category |  |  |  |  |
| Children (5-15 years) | 595 | 81% | 90 | 77% |
| Adolescents (16-19 years) | 140 | 19% | 54 | 23% |
| Sex |  |  |  |  |
| Girls | 341 | 46% | 71 | 50% |
| Boys | 394 | 54% | 73 | 50% |
| Migration background |  |  |  |  |
| No | 552 | 75% | 118 | 76% |
| Yes | 183 | 25% | 26 | 24% |
| Language |  |  |  |  |
| German | 516 | 70% | 105 | 69% |
| French | 185 | 25% | 33 | 26% |
| Italian | 34 | 5% | 6 | 4% |
| Parental education |  |  |  |  |
| Primary education | 45 | 6% | 2 | 1% |
| Secondary education | 242 | 33% | 44 | 30% |
| Tertiary education | 428 | 58% | 98 | 68% |
| Missing | 20 | 3% | 0 | 0% |
| Siblings |  |  |  |  |
| No | 88 | 12% |  |  |
| Yes | 630 | 86% |  |  |
| Missing | 17 | 2% |  |  |
| **Cancer-related characteristics<sup>d</sup>** | **N** | **%** |  |  |
| ICCC3 main group |  |  |  |  |
| Leukemias | 299 | 41% |  |  |
| Lymphomas | 60 | 8% |  |  |
| CNS | 123 | 17% |  |  |
| Neuroblastoma | 54 | 7% |  |  |
| Retinoblastoma | 29 | 4% |  |  |
| Renal tumor | 48 | 7% |  |  |
| Hepatic tumor | 6 | 1% |  |  |
| Malignant bone tumors | 17 | 2% |  |  |
| Soft tissue sarcomas | 52 | 7% |  |  |
| Germ cell tumors | 16 | 2% |  |  |
| Other | 5 | 1% |  |  |
| Langerhans cell histiocytosis | 26 | 4% |  |  |
| Age at diagnosis, mean (SD) | 4.9 (3.5) |  |  |  |
| median (IQR) | 4.2 [2.1 – 7.2] |  |  |  |
| Time since diagnosis, mean (SD) | 8.3 (1.9) |  |  |  |
| median (IQR) | 8.2 [6.9 – 9.4] |  |  |  |
| Year of diagnosis, mean (SD) | 2010 (3.2) |  |  |  |
| median (IQR) | 2010 [2005–2015] |  |  |  |
| Radiotherapy | 137 | 19% |  |  |
| Cranial radiotherapy | 68 | 9% |  |  |
| Chemotherapy | 588 | 80% |  |  |
| HSCT | 69 | 9% |  |  |
| Autologous | 30 | 4% |  |  |
| Allogeneic | 39 | 5% |  |  |
| Relapse | 81 | 11% |  |  |

<sup>a</sup> For siblings, the percentages are standardized to survivors based on sex, age at study, migration background, and language. The N is the original value. <sup>b</sup> Calculated based upon weighted t-test for continuous variables and weighted chi- square tests for categorical variables. <sup>c</sup> Not applicable since siblings were standardized based upon these variables to match the CCS. <sup>d</sup> No missing values for cancer-related characteristics.
ICCC3 = International Classification of Childhood Cancer, third edition; HSCT = hematopoietic stem cell transplantation

### Prevalence of dental problems in CCS and comparison with siblings

CCS and siblings reported having at least one dental problem with similar frequency, but the type of dental problems differed (*Figure 1* and *Supplementary Table 2*). Eight percent of CCS reported having hypodontia (95% confidence interval (CI) 6-10), 11% microdontia (95% CI 8-13), and 8% (95% CI 6-10) enamel hypoplasia. Cavities were prevalent in 39% of CCS (95% CI 35-43) and tooth loss due to cavities or gum disease in 6% (95% CI 4-7). Thirty-four percent (95% CI 30-37) of CCS reported gum problems during treatment, and 14% (95% CI 11-17) after treatment. When we stratified the analysis by age of diagnosis, we found that compared to cancer patients diagnosed at 5 years or older, cancer patients diagnosed before the age of 5 years had higher prevalence of hypodontia (10% vs. 4%) and microdontia (14% vs. 5%). Gum problems during treatment were more prevalent in cancer patients diagnosed at 5 years or older (39%) compared to those diagnosed before the age of 5 years (30%) (*Supplementary Table 3*). For an overview of prevalence rates in siblings without survey weights and dental problems reported in open questions, see *Supplementary Table 4*.

**FIGURE 1.**
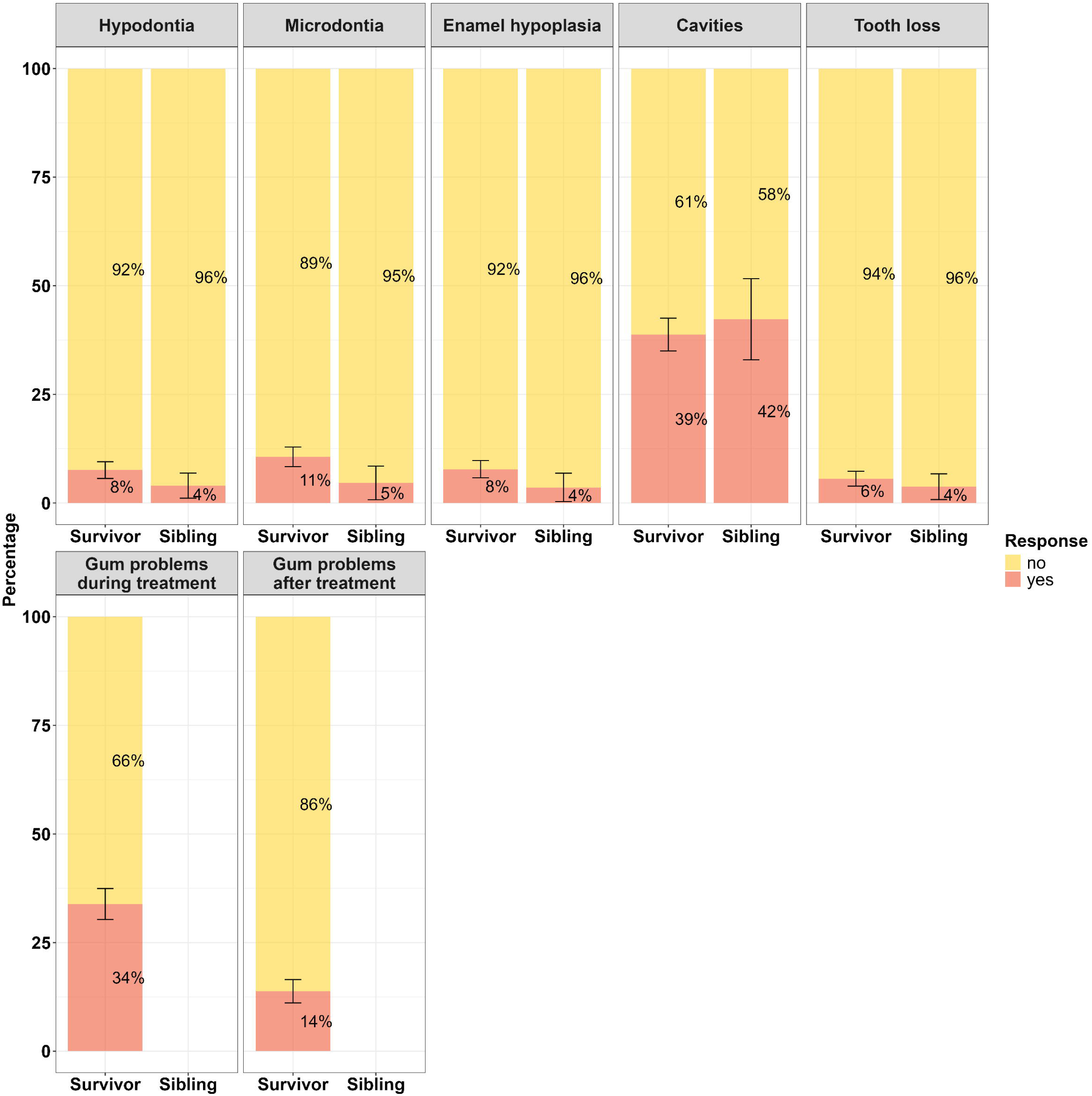
Prevalence of dental problems in childhood cancer survivors and siblings. Please note: Only CCS reported gum problems during and after treatment. Percentages are based upon the number of respondents for each dental problem reported in *Table 2*. For siblings, we report weighted percentages.

**TABLE 2.**
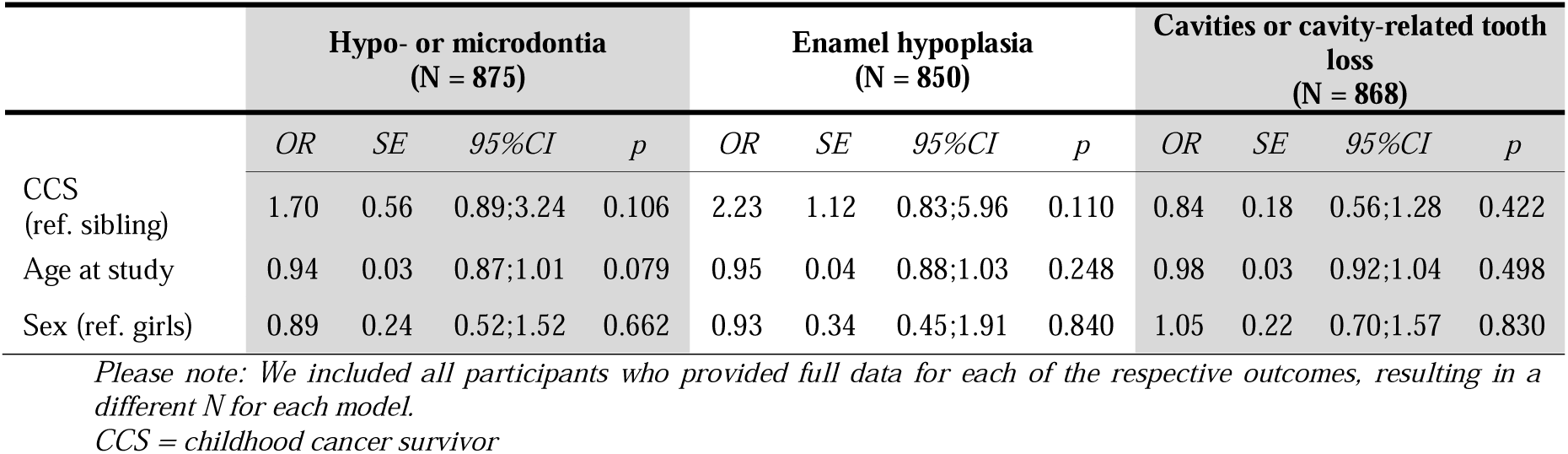
Dental problems in childhood cancer survivors compared to siblings in multivariable logistic regressions.

|  | Hypo- or microdontia<br>(N = 875) |  |  |  | Enamel hypoplasia<br>(N = 850) |  |  |  | Cavities or cavity-related tooth<br>loss<br>(N = 868) |  |  |  |
| --- | --- | --- | --- | --- | --- | --- | --- | --- | --- | --- | --- | --- |
|  | OR | SE | 95%CI | p | OR | SE | 95%CI | p | OR | SE | 95%CI | p |
| CCS<br>(ref. sibling) | 1.70 | 0.56 | 0.89;3.24 | 0.106 | 2.23 | 1.12 | 0.83;5.96 | 0.110 | 0.84 | 0.18 | 0.56;1.28 | 0.422 |
| Age at study | 0.94 | 0.03 | 0.87;1.01 | 0.079 | 0.95 | 0.04 | 0.88;1.03 | 0.248 | 0.98 | 0.03 | 0.92;1.04 | 0.498 |
| Sex (ref. girls) | 0.89 | 0.24 | 0.52;1.52 | 0.662 | 0.93 | 0.34 | 0.45;1.91 | 0.840 | 1.05 | 0.22 | 0.70;1.57 | 0.830 |

Please note: We included all participants who provided full data for each of the respective outcomes, resulting in a different N for each model.
CCS = childhood cancer survivor

After controlling for age and sex, we found a trend towards an increased risk for hypo- or microdontia (OR=1.70, 95% CI 0.89-3.24) and enamel hypoplasia (OR=2.23, 95% CI 0.83-5.96) in CCS compared to siblings. Cavities and tooth loss were similar between CCS and siblings (OR=0.84, 95% CI 0.56-1.28) (see *Table 2*).

Of all CCS reporting at least one dental problem (N = 329), 20% reported problems with health care insurance covering associated costs. Insurance problems varied by dental problem (see *Supplementary Table 5*).

### Cancer-related risk factors of dental problems in CCS

After controlling for age at diagnosis, time since diagnosis, and relapse, chemotherapy treatment was consistently associated with dental problems. This included enamel hypoplasia (OR=3.08, 95% CI 1.22-10.40), cavities or cavity-related tooth loss (OR=1.52, 95% CI 1.01-2.31) as well as gum problems during (OR=22.97, 95% CI 9.37-76.23) and after cancer treatment (OR=4.62, 95% CI 1.98-13.49). We found similar associations between HSCT and dental problems. Specifically, cancer patients who underwent HSCT had an increased risk for hypo- or microdontia (OR = 5.37, 95%CI 2.64-10.69), cavities or cavity-related tooth loss (OR = 2.10, 95%CI 1.24-3.57), and gum problems during treatment (OR = 2.03, 95%CI 1.16-3.60) compared to CCS without HSCT. Cranial radiation was unrelated to all dental problems (see *Table 3*).

**TABLE 3.** Cancer-related risk factors of dental problems in childhood cancer survivors in multivariable logistic regression.

|  | Hypo- or microdontia<br>(N = 725) |  |  |  | Enamel hypoplasia<br>(N = 706) |  |  |  | Cavities or<br>cavity-related tooth loss<br>(N = 724) |  |  |  |
| --- | --- | --- | --- | --- | --- | --- | --- | --- | --- | --- | --- | --- |
|  | OR | SE | 95%CI | p | OR | SE | 95%CI | p | OR | SE | 95%CI | p |
| Cranial radiation<br>(ref. no) | 0.85 | 0.46 | 0.26;2.25 | 0.762 | 0.80 | 0.42 | 0.25;2.07 | 0.676 | 0.88 | 0.25 | 0.50;1.51 | 0.637 |
| Chemotherapy (ref.<br>no) | 1.59 | 0.73 | 0.70;4.32 | 0.309 | 3.08 | 1.64 | 1.22;10.40 | **0.035** | 1.52 | 0.32 | 1.01;2.31 | **0.049** |
| HSCT (ref. no) | 5.37 | 1.91 | 2.64;10.69 | **<0.001** | 1.74 | 0.70 | 0.75;3.70 | 0.171 | 2.10 | 0.56 | 1.24;3.57 | **0.006** |
| Age at diagnosis | 0.85 | 0.04 | 0.76;0.94 | **0.002** | 0.92 | 0.04 | 0.83;1.00 | 0.069 | 1.04 | 0.02 | 0.99;1.09 | 0.094 |
| Time since<br>diagnosis | 1.08 | 0.07 | 0.94;1.23 | 0.248 | 1.02 | 0.08 | 0.87;1.17 | 0.834 | 1.12 | 0.05 | 1.04;1.22 | **0.005** |
| Relapse (ref. no) | 1.11 | 0.48 | 0.45;2.49 | 0.814 | 2.23 | 0.86 | 1.01;4.63 | **0.038** | 0.62 | 0.17 | 0.35;1.05 | 0.082 |
|  | Gum problems during treatment<br>(N = 676) |  |  |  | Gum problems after treatment<br>(N = 637) |  |  |  |  |  |  |  |
|  | OR | SE | 95%CI | p | OR | SE | 95%CI | p |  |  |  |  |
| Cranial radiation<br>(ref. no) | 1.52 | 0.45 | 0.84;2.72 | 0.163 | 1.46 | 0.52 | 0.70;2.87 | 0.287 |  |  |  |  |
| Chemotherapy (ref.<br>no) | 22.97 | 11.97 | 9.37;76.23 | **<0.001** | 4.62 | 2.21 | 1.98;13.49 | **0.001** |  |  |  |  |
| HSCT (ref. no) | 2.03 | 0.59 | 1.16;3.60 | **0.014** | 1.65 | 0.57 | 0.82;3.18 | 0.147 |  |  |  |  |
| Age at diagnosis | 1.11 | 0.03 | 1.06;1.18 | **<0.001** | 1.09 | 0.04 | 1.02;1.17 | **0.010** |  |  |  |  |
| Time since<br>diagnosis | 1.01 | 0.05 | 0.92;1.12 | 0.777 | 1.13 | 0.07 | 1.00;1.28 | **0.041** |  |  |  |  |
| Relapse (ref. no) | 1.04 | 0.31 | 0.57;1.85 | 0.907 | 1.45 | 0.52 | 0.70;2.85 | 0.301 |  |  |  |  |

Please note: We included all participants who provided full data for each of the respective outcomes, resulting in a different N for each model.
HSCT = hematopoietic stem cell transplantation

Age stratified analyses showed that associations between cancer-related risk factors and dental problems differed by age group (see *Table 4*). Cancer patients diagnosed before the age of 5 years had an increased risk for hypo- or microdontia when treated with chemotherapy (OR=5.89, 95% CI 2.08-24.80) or HSCT (OR=2.76, 95% CI 1.34-5.58). They were also at higher risk for cavities or cavity-related tooth loss when treated with HSCT (OR=3.25, 95% CI 1.64-6.59). We did not observe those associations in cancer patients diagnosed at the age of 5 years or older. All other treatment-related risk factors were similarly related to dental problems across age diagnosis groups.

**TABLE 4.**
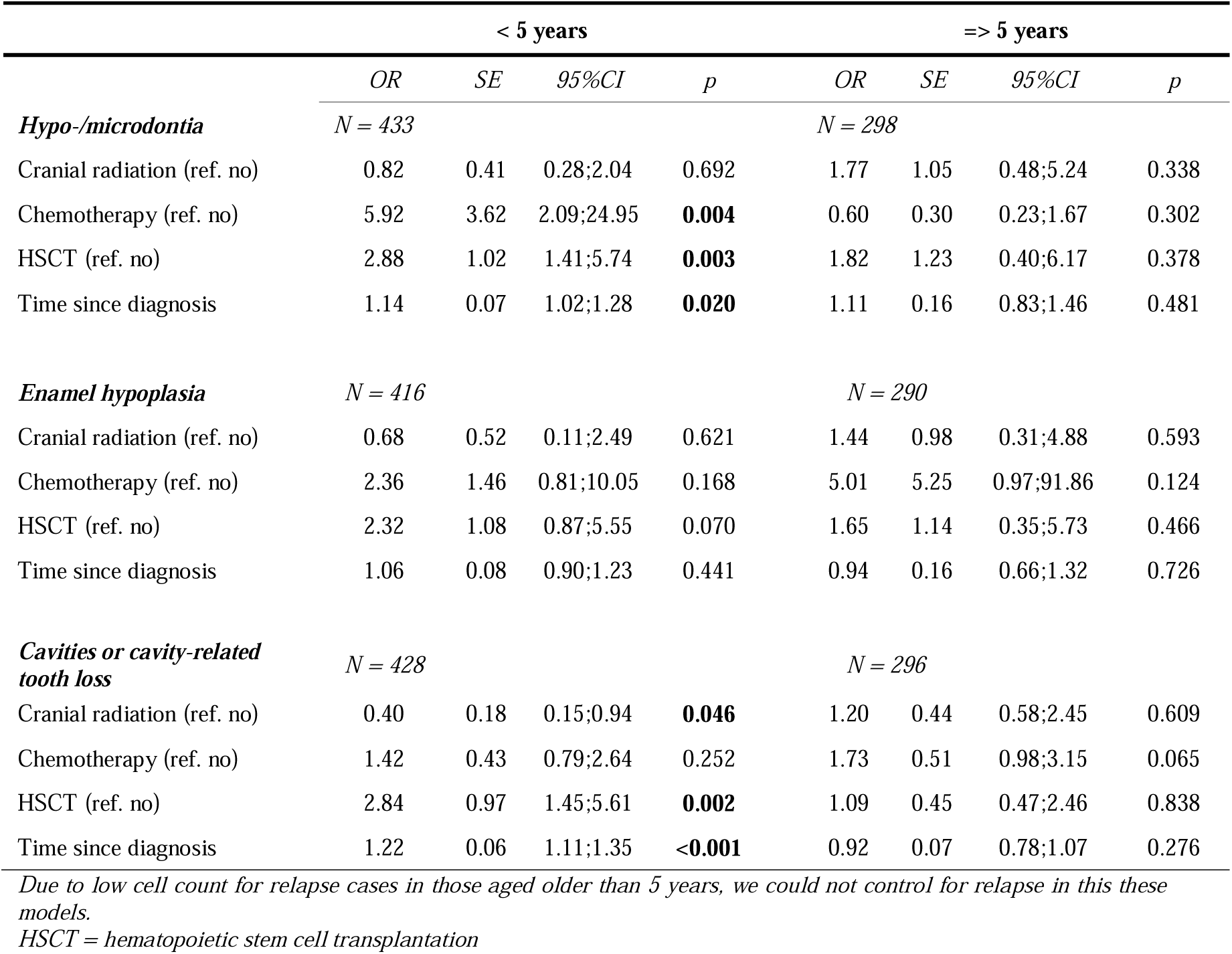
Age-stratified analysis for cancer-related characteristics and dental problems in childhood cancer survivors.

### Sensitivity analysis

In imputed data analysis, we included *N* = 907 study participants (*N* = 763 CCS, *N* = 144 siblings). Analyses yielded the same conclusions as complete-case analysis; in fact, most associations became stronger and confidence intervals smaller (see *Supplementary Table 6* and *Supplementary Table 7*).

## Discussion

This national survey in Switzerland found that CCS were at higher risk for hypo- or microdontia and enamel hypoplasia than their siblings, while cavities and cavity-related tooth loss did not differ between CCS and siblings. Chemotherapy and HSCT were consistently related to dental problems. Cancer patients diagnosed before the age of 5 and treated with chemotherapy or HSCT were particularly susceptible to hypo- or microdontia, as well as cavities or cavity-related tooth loss.

The most prevalent dental problem in CCS was cavities. It is difficult to compare prevalence rates for cavities and cavity-related tooth loss to other studies with pediatric CCS since those studies used the composite Decayed, Missing, Filled Teeth (DMFT) score obtained through dental examinations.^8,28^ Existing population-based or cohort studies using questionnaires with adult CCS showed that similar to our study, cavities were the most prevalent problem across all dental problems investigated.^12,17^ While in our study, prevalence was 8% for hypodontia and 11% for each microdontia and enamel hypoplasia, other studies with CCS of similar age indicate higher prevalence for microdontia (16-31%), hypodontia (11-36%), and enamel hypoplasia (12-32%).^16,28–31^ These differences are likely due to those studies assessing dental problem using dental examinations and clinical methods such as radiographic imaging. When comparing to CCS questionnaire studies in adults, prevalence rates were similar for microdontia (9-14%), hypodontia (8-10%), and enamel hypoplasia (12-17%).^12,17^ Higher prevalence of hypo- and microdontia in CCS diagnosed in early childhood was in line with previous findings,^29,30^ and mirrors young children’s susceptibility to dental late effects of cancer treatment due to being in a crucial dentition phase.^9^

In line with our results, studies from clinical examination in pediatric CCS ^16^ and in population-based or cohort questionnaire studies with adult CCS ^12,17^ found a higher risk for hypo- or microdontia and enamel hypoplasia in CCS than in controls. Population-based and cohort questionnaire studies with adult CCS also showed that cavity risk is similar in CCS and siblings.^12,17^ Similar cavity risk could be due to increased awareness regarding the importance of dental care during cancer treatment, with clinical practitioners taking preventive measures against cavity-related risk factors induced by cancer treatment, such as xerostomia. Another potential explanation is the strong impact of diet and dental hygiene on cavities,^32,33^ which is likely similar within families (i.e., CCS and the sibling control group), masking the impact of cancer treatment on dental health.

Among cancer-related risk factors, chemotherapy was linked to higher risks of enamel hypoplasia, cavities or tooth loss due to cavities, and gum problems both during and after treatment. Previous studies utilizing clinical examinations have reported comparable findings regarding the effects of chemotherapy.^8^ HSCT was associated with increased risks of hypo- or microdontia, cavities or tooth loss due to cavities, and gum problems during treatment Our results are similar to findings from a review summarizing results of ten studies that CCS who underwent HSCT had an increased risk for micro- and hypodontia, while findings regarding the impact of HSCT on cavities are conflicting.^34^ We found that the risk for dental problems was high when chemotherapy or HSCT was administered at or before the age of 5 years. This demonstrates the profound impact of cancer treatment on dentition in the early years when crown and root formation are occuring.^11^ Cranial radiation was unrelated to all dental problems in both overall and age-stratified analysis, which contrasts previous findings.^10,11^ There are several potential explanations for this finding. The radiation location in the head or neck-area may not have involved the salivary glands or tooth buds, or the radiation dose could have been too low to cause damage. In Switzerland, proton instead of photon radiotherapy has been increasingly used, which may cause fewer dental problems.^35–37^ Further investigations into the underlying reasons are necessary.

With one out of five survivors reporting issues with health insurance coverage for dental problems, our study implies that assessing dental health before cancer treatment is crucial to secure insurance coverage for potential post-treatment dental problems in healthcare systems where health insurance covers dental costs only if they are a sequalae of a disease (such as in Switzerland). Healthcare providers are essential to support survivors and their parents in obtaining health insurance coverage and to help CCS monitoring and maintaining dental health. This includes encouraging annual dental check-ups following COG guidelines^11^ to treat potential conditions and avoid further damage regarding development of the jaw and other teeth resulting from those conditions. Fostering dental hygiene habits such as brushing, flossing, and maintaining a low-sugar diet can further reduce the risk of cavities. Supporting older children and adolescent survivors of childhood cancers in avoiding behaviors like smoking and excessive alcohol consumption can also contribute to better long-term dental health.^38,39^

Our population-based, representative study adds evidence to the field of dental late effects by providing results from pediatric CCS between 5-19 years, reducing recall bias, providing insights into dental problems of CCS outside North America, and comparing the results to a control group having a similar context as CCS. Previous studies covered only adult CCS or were clinical studies with smaller and selective pediatric populations. We combined patient-reported information with systematically coded clinical information from the ChCR. Our study uses self-reported information about dental problems, which may lead to underestimation of dental late effects. The relatively small number of siblings in the control group may have introduced selection bias. Although we had a large cohort, some dental problems were rare, leading to limited statistical power and bearing the risk that we may have missed associations when comparing the risk for dental problems between CCS and siblings. When we imputed missing data, the associations became stronger and confidence intervals smaller, strengthening confidence in our conclusions. Finally, we did not have detailed treatment information, such as the use of alkylating agents or dental status at the time of treatment, which can modify the effect of cancer-treatment on dental late effects.^11^

In conclusion, we found that child and adolescent survivors of childhood cancer have an increased risk for hypodontia, microdontia, and enamel hypoplasia compared to their siblings. Healthcare providers should be aware of the increased risk of dental problems among child and adolescent survivors of childhood cancers, especially if patients were treated with chemotherapy or HSCT before the age of five years.

## Acknowledgements

We thank all survivors for participating in our study, the study team of the Childhood Cancer Research Group, the data managers of the Swiss Paediatric Oncology Group, and the team of the Swiss Childhood Cancer Registry.

## Data availability statement

The data that support the information of this manuscript were accessed on secured servers of the Institute of Social and Preventive Medicine at the University of Bern. Individual-level, fully anonymized, sensitive data can only be made available for researchers who fulfil the respective legal requirements. Requests of data from the Childhood Cancer Registry must be directed to the Childhood Cancer Registry of Switzerland (https://www.childhoodcancerregistry.ch). Requests of data from the Swiss Childhood Cancer Survivor Study (SCCSS) should be communicated to the study lead Claudia E. Kuehni.

## Abbreviations

CCS: childhood cancer survivors
HSCT: hematopoietic stem cell transplantation
ChCR: Swiss Childhood Cancer Registry
COG: Children’s Oncology Group
ICCC3: International Classification of Childhood Cancer, third edition

## References

1. Schindler M, Belle FN, Grotzer MA, Von Der Weid NX, Kuehni CE. Childhood cancer survival in Switzerland (1976–2013): Timeltrends and predictors. Int J Cancer. 2017;140(1):62–74. doi:10.1002/ijc.30434

2. Bhakta N, Liu Q, Ness KK, et al. The cumulative burden of surviving childhood cancer: an initial report from the St Jude Lifetime Cohort Study (SJLIFE). Lancet. 2017;390(10112):2569–2582. doi:10.1016/s0140-6736(17)31610-0

3. Hammoud RA, Liu Q, Dixon SB, et al. The burden of cardiovascular disease and risk for subsequent major adverse cardiovascular events in survivors of childhood cancer: a prospective, longitudinal analysis from the St Jude Lifetime Cohort Study. Lancet Oncol. 2024;25(6):811–822. doi:10.1016/s1470-2045(24)00157-8

4. Christen S, Roser K, Mader L, et al. Incidence and prevalence of musculoskeletal health conditions in survivors of childhood and adolescent cancers: A report from the Swiss childhood cancer survivor study. Cancer Medicine. 2024;13(8):e7204. doi:10.1002/cam4.7204

5. Weiss A, Sommer G, Kasteler R, et al. Long-term auditory complications after childhood cancer: A report from the Swiss Childhood Cancer Survivor Study. Pediatr Blood Cancer. 2017;64(2):364–373. doi:10.1002/pbc.26212

6. Kasteler R, Weiss A, Schindler M, et al. Longlterm pulmonary disease among Swiss childhood cancer survivors. Pediatr Blood Cancer. 2018;65(1):e26749. doi:10.1002/pbc.26749

7. Whelan KF, Stratton K, Kawashima T, et al. Ocular late effects in childhood and adolescent cancer survivors: A report from the childhood cancer survivor study. Pediatr Blood Cancer. 2010;54(1):103–109. doi:10.1002/pbc.22277

8. Busenhart DM, Erb J, Rigakos G, Eliades T, Papageorgiou SN. Adverse effects of chemotherapy on the teeth and surrounding tissues of children with cancer: A systematic review with meta-analysis. Oral Oncol. 2018;83:64–72. doi:10.1016/j.oraloncology.2018.06.001

9. Smith BH. Standards of human tooth formation and dental age assessment. Wiley-Liss Inc.; 1991.

10. Milgrom SA, van Luijk P, Pino R, et al. Salivary and Dental Complications in Childhood Cancer Survivors Treated With Radiation Therapy to the Head and Neck: A PENTEC Comprehensive Review. Int J Radiat Oncol Biol Phys. 2024;119(2):467–481. doi:10.1016/j.ijrobp.2021.04.023

11. Effinger KE, Migliorati CA, Hudson MM, et al. Oral and dental late effects in survivors of childhood cancer: a Children’s Oncology Group report. Support Care Cancer. 2014;22(7):2009–2019. doi:10.1007/s00520-014-2260-x

12. Kaste SC, Goodman P, Leisenring W, et al. Impact of radiation and chemotherapy on risk of dental abnormalities. Cancer. 2009;115(24):5817–5827. doi:10.1002/cncr.24670

13. Kaste SC, Hopkins KP, Jenkins JJI. Abnormal odontogenesis in children treated with radiation and chemotherapy: imaging findings. Am J Roentgenol. 1994;162(6):1407–1411. doi:10.2214/ajr.162.6.8192008

14. Imanguli MM, Atkinson JC, Mitchell SA, et al. Salivary Gland Involvement in Chronic Graft-Versus-Host Disease: Prevalence, Clinical Significance, and Recommendations for Evaluation. Biol Blood Marrow Transplant. 2010;16(10):1362–1369. doi:10.1016/j.bbmt.2010.03.023

15. Dahllöf G, Wondimu B, Barr-Agholme M, Garming-Legert K, Remberger M, Ringdén O. Xerostomia in children and adolescents after stem cell transplantation conditioned with total body irradiation or busulfan. Oral Oncol. 2011;47(9):915–919. doi:10.1016/j.oraloncology.2011.06.509

16. Pombo Lopes J, Rodrigues I, Machado V, Botelho J, Bandeira Lopes L. Chemotherapy and Radiotherapy Long-Term Adverse Effects on Oral Health of Childhood Cancer Survivors: A Systematic Review and Meta-Analysis. Cancers. 2023;16(1):110. doi:10.3390/cancers16010110

17. Patni T, Lee C-T, Li Y, et al. Factors for poor oral health in long-term childhood cancer survivors. BMC Oral Health. 2023;23(1):73. doi:10.1186/s12903-023-02762-0

18. Maes L, Vereecken C, Vanobbergen J, Honkala S. Tooth brushing and social characteristics of families in 32 countries. Int Dent J. 2006;56(3):159–167. 10.1111/j.1875-595X.2006.tb00089.x

19. Zaborskis A, Milciuviene S, Narbutaite J, Bendoraitiene E, Kavaliauskiene A. Caries experience and oral health behaviour among 11 −13-year-olds: an ecological study of data from 27 European countries, Israel, Canada and USA. Community Dent Health. Jun 2010;27(2):102–8.

20. Kuehni CE, Rueegg CS, Michel G, et al. Cohort Profile: The Swiss Childhood Cancer Survivor Study. Int J Epidemiol. 2012;41(6):1553–1564. doi:10.1093/ije/dyr142

21. Michel G, von der Weid NX, Zwahlen M, Adam M, Rebholz C, Kuehni CE. The Swiss Childhood Cancer Registry: rationale, organisation and results for the years 2001-2005. Swiss Med Wkly. 2007;137(3536):502–509.

22. Statistik Bf. Bevölkerung nach Migrationsstatus [Population and migration status]. Accessed 17 April, 2024, https://rb.gy/nf754k

23. Statistik Bf. Bildungssystem [Education system]. Accessed 19 April, 2024, https://www.bfs.admin.ch/bfs/de/home/statistiken/bildung-wissenschaft/bildungssystem.assetdetail.223674.html

24. SteliarovalFoucher E, Stiller C, Lacour B, Kaatsch P. International classification of childhood cancer. Cancer. 2005;103(7):1457–1467.

25. Gianinazzi ME, Rueegg CS, Von Der Weid NX, Niggli FK, Kuehni CE, Michel G. Mental health-care utilization in survivors of childhood cancer and siblings: the Swiss childhood cancer survivor study. Support Care Cancer. 2014;22(2):339–349. doi:10.1007/s00520-013-1976-3

26. Van Buuren S, Groothuis-Oudshoorn K. mice: Multivariate imputation by chained equations in R. Journal of Statistical Software. 2011;45:1–67.

27. Carpenter JR, Bartlett JW, Morris TP, Wood AM, Quartagno M, Kenward MG. Multiple imputation and its application. John Wiley & Sons; 2023.

28. Defabianis P, Bocca N, Romano F. Prevalence and association of dental anomalies and tooth decay in Italian childhood cancer survivors. J Clin Pediatr Dent. 2023;doi:10.22514/jocpd.2023.056

29. Halperson E, Matalon V, Goldstein G, et al. The prevalence of dental developmental anomalies among childhood cancer survivors according to types of anticancer treatment. Sci Rep. 2022;12(1)doi:10.1038/s41598-022-08266-1

30. Kang C-M, Hahn SM, Kim HS, et al. Clinical Risk Factors Influencing Dental Developmental Disturbances in Childhood Cancer Survivors. Cancer Res Treat. 2018;50(3):926–935. doi:10.4143/crt.2017.296

31. Quispe RA, Rodrigues ACC, Buaes AMG, Capelozza ALA, Rubira CMF, Santos PSdS. A case-control study of dental abnormalities and dental maturity in childhood cancer survivors. Oral Surg Oral Med Oral Radiol. 2019/11/01/ 2019;128(5):498–507.e3. doi:10.1016/j.oooo.2019.07.005

32. Moynihan P, Petersen PE. Diet, nutrition and the prevention of dental diseases. Public Health Nutr. 2004;7(1a):201–226. doi:10.1079/phn2003589

33. Kumar S, Tadakamadla J, Johnson NW. Effect of Toothbrushing Frequency on Incidence and Increment of Dental Caries:A Systematic Review and Meta-Analysis. J Dent Res. 2016;95(11):1230–1236. doi:10.1177/0022034516655315

34. Gawade PL, Hudson MM, Kaste SC, et al. A systematic review of dental late effects in survivors of childhood cancer. Pediatr Blood Cancer. 2014;61(3):407–416. doi:10.1002/pbc.24842

35. Martinsson U, Svärd A-M, Witt Nyström P, et al. Complications after proton radiotherapy in children, focusing on severe late complications. A complete Swedish cohort 2008–2019. Acta Oncol. 2023;62(10):1348–1356. doi:10.1080/0284186x.2023.2260946

36. Taylor CL, Price JM. The Tooth Hurts: Dental Health After Radiation Therapy for Head and Neck Cancer. Int J Radiat Oncol Biol Phys. 2022;113(2):331–334. doi:10.1016/j.ijrobp.2022.01.005

37. Grant SR, Grosshans DR, Bilton SD, et al. Proton versus conventional radiotherapy for pediatric salivary gland tumors: Acute toxicity and dosimetric characteristics. Radiother Oncol. 2015;116(2):309–315. doi:10.1016/j.radonc.2015.07.022

38. Dentistry AAoP. Dental Management of Pediatric Patients Receiving Immunosuppressive Therapy and/or Head and Neck Radiation. The Reference Manual of Pediatric Dentistry. 2023.

39. Hong CH, daFonseca M. Considerations in the Pediatric Population with Cancer. Dent Clin North Am. 2008;52(1):155–181. doi:10.1016/j.cden.2007.10.001

