## Supplementary File for "Dental health of childhood cancer survivors – a report from the Swiss Childhood Cancer Survivor Study (SCCSS)"

Carina Nigg, Corinne Matti, Philippa Jörger, André von Bueren, Cornelia Filippi, Tamara Diesch-Furlanetto, Zuzana Tomášiková, Claudia E Kuehni, Grit Sommer

Pediatric Blood & Cancer

**Corresponding author:**

Prof. Claudia E. Kuehni, MD, MSc

Institute of Social and Preventive Medicine

University of Bern

Mittelstrasse 43

3012 Bern

Switzerland

**Table of content**

### **Supplementary literature summary**

| **Author,**  **year** | **Study type** | **Country** | **Sample size** | **Age (mean or median)** | **Dental health assessment** | **Prevalence of dental problems in CCS and control group (if applicable)** | **Risk factors of dental problems** |
| --- | --- | --- | --- | --- | --- | --- | --- |
| **Systematic reviews & meta-analysis** | | | | | | | |
| Busenhart et al., 2018^1^ | Systematic review with meta-analysis of clinical case-control studies | 13 countries | 16 articles with at least 2,315 CCS treated with chemotherapy but not radiotherapy (9 studies post-therapy, 3 studies during therapy, 3 studies did not report)  CG: Children with or without cancer not treated with chemotherapy | Age at study: < 18 years  Age at dx: mean = 6,6 years | Clinical |  | ***Microdontia (4 studies)***  Chemotherapy group: RR = 12.41, 95%CI 3.05–50.60  ***Tooth agenesis (5 studies)***  Chemotherapy group: RR = 2.47, 95%CI 1.30-4.71  ***Enamel hypoplasia (2 studies)***  Chemotherapy group: RR=3.08, 95%CI 1.13-8.37  ***Cavities (DMFT; 3 studies)***  Chemotherapy group: mean difference =3.07 teeth; 95%CI 2.26–3.88  ***Missing teeth index (2 studies)***  Chemotherapy group: mean difference = −0.56 teeth, 95%CI −1.06 to −0.07  ***Gingival index (3 studies)***  Chemotherapy group: mean difference = 0.38 units, 95%CI 0.16–0.61 |
| Popom Lopes et al., 2023^2^ | Systematic review with meta-analysis including studies of any design | Various | 35 articles with 2625 CCS included in review; 22 articles included in meta-analysis  CG: 1136 healthy children | Age at dx: 3-18 years  Age at study: NR | Clinical | *Microdontia (14 studies)*  16%, 95%CI 9-24  *Tooth agenesis (10 studies)*  36%, 95%CI 27-45  *Hypodontia*  13%, 95%CI 5-23  *Enamel hypoplasia*  32%, 95%CI 21-45% | ***Microdontia (7 studies)***  CCS compared to healthy children: OR = 9.49, 95%CI 3.13-28.70  ***Tooth agenesis (8 studies)***  CCS compared to healthy children: OR = 3.50, 95%CI 1.98-6.16  ***Enamel hypoplasia (7 studies)***  CCS compared to healthy children: OR = 1.95, 95%CI 1.32-2.88 |
| Seremedi et al., 2019^3^ | Systematic review and meta-analysis of retrospective non-randomized studies | Various | 16 articles with 1300 CCS post-treatment with a combination of chemotherapy and cranial radiation (head and neck area; TBI) up to the age of 12 years  CG: Not required | Age at study: 11-15 years (range of reported mean/median)  Age at dx: 2.7-6.9 years (range of reported mean/median) | NR |  | *Microdontia*   - **< 4.5 years at diagnosis (vs. ≥ 4.5 years)**: OR = 7.0, p < 0.05 - **TBI (vs. cranial radiation)**: OR = 2.3, p < 0.05 - HSCT (vs. no HSCT): OR = 0.4, p > 0.05   *Malformed teeth*   - < 4.5 years at diagnosis (vs. ≥ 4.5 years): OR = 0.8, p > 0.05 - TBI (vs. cranial radiation): OR = 1.2, p > 0.05 - HSCT (vs. no HSCT): OR = 0.5, p > 0.05   *Hypodontia*   - < 4.5 years at diagnosis (vs. ≥ 4.5 years): OR = 1.1, p > 0.05 - TBI (vs. cranial radiation): OR = 0.7, p > 0.05 - HSCT (vs. no HSCT): OR = 0.8, p > 0.05   *Tooth agenesis*   - < 4.5 years at diagnosis (vs. ≥ 4.5 years): OR = 4.0, p > 0.05 - TBI (vs. cranial radiation): OR = 2.5, p > 0.05 - HSCT (vs. no HSCT): OR = 0.3, p > 0.05   *Enamel hypoplasia*   - < 4.5 years at diagnosis (vs. ≥ 4.5 years): OR = 4.7, p > 0.05 - TBI (vs. cranial radiation): OR = 0.3, p > 0.05 - HSCT (vs. no HSCT): OR = 0.7, p > 0.05 |
| **Single studies with pediatric cancer survivors** | | | | | | | |
| Defabianis et al, 2023^4^ | Retrospective | Italy | 88 CCS > 2 years in remission treated with chemotherapy and/or radiotherapy  No CG | Age at study: 11.4 ± 4.2 years  Age at dx: 5.1 ± 3.1 years | Intra-oral examination, orthopantomography | *Microdontia*  28.4%  *Tooth agenesis*  28.4%  *Enamel hypoplasia*  11.6-27.9% depending on degree  *Cavities (DMFT)*  mean = 3.5 ± 3 | *Microdontia*   - **Age at treatment** (p < 0.001)   < 5 years: 46.9% ≥ 5 years: 5.1%   - **Type of cancer therapy** (p < 0.01)   Chemotherapy: 43.9%  Radiotherapy: 7.7%  Chemo- + radiotherapy: 17.6%  *Tooth agenesis*   - **Age at treatment** (p < 0.001)   < 5 years: 46.9% ≥ 5 years: 5.1%   - Type of cancer therapy (p > 0.05)   Chemotherapy: 34.1%  Radiotherapy: 15.4%  Chemo- + radiotherapy: 26.5%  *Enamel hypoplasia*  No association with age of treatment or type of cancer therapy for any enamel hypoplasia degree  *Cavities/tooth loss: DMFT score*   - Age at treatment (p > 0.05)   < 5 years: 3.0 ± 2.6 ≥ 5 years: 4.1 ± 3.3   - Type of cancer therapy (p > 0.05)   Chemotherapy: 3.3 ± 2.7  Radiotherapy: 3.4 ± 3.5  Chemo- + radiotherapy: 3.9 ± 3.3 |
| Halperson et al., 2022^5^ | Retrospective | Israel | 121 CCS in survivorship care  No CG | Age at study: 15.9 years  Age at dx: 7.1 years | Dental examination | Overall  *Microdontia:*  17%  *Hypodontia:*  11%  *Hypocalcification or enamel hypoplasia:*  17%  Age at treatment  ***Microdontia***   - ≤ 6 years at treatment: 33% - > 6 years at treatment: 7%   p = 0.002  ***Hypodontia***   - ≤ 6 years at treatment: 20% - > 6 years at treatment: 2%   p = 0.007  ***Total number of malformed teeth***   - ≤ 6 years at treatment: 4.15 ± 6.85 - > 6 years at treatment: 1.67 ± 3.82   p = 0.013  *Hypocalcification or hypoplasia*   - ≤ 6 years at treatment: 15% - > 6 years at treatment: 23%   p = 0.269  *Cavities (DMFT)*   - ≤ 6 years at treatment: 6.07 ± 6.49 - > 6 years at treatment: 6.02 ± 4.70   p = 0.483 | **Prevalence by treatment (no p-values reported)**  *Microdontia:*   - Chemotherapy only: 19% - Bone marrow transplant: 19% - Cranial radiation (head/neck): 20%   *Hypodontia*   - Chemotherapy only: 11% - Bone marrow transplant: 11% - Cranial radiation (head/neck): 13%   *Hypocalcification or hypoplasia*   - Chemotherapy only: 13% - Bone marrow transplant: 14% - Cranial radiation (head/neck): 33%   *Cavities (DMFT; mean ± SD):*   - Chemotherapy only: 5.93 ± 5.73 - Bone marrow transplant: 6.67 ± 6.85 - Cranial radiation (head/neck): 7.93 ± 5.46 |
| Kang et al, 2018^6^ | Retrospective | Korea | 196 CCs > 2 years in remission  No CG | Age at study: 15.6 years  Age at dx: 4.7 years | Dental examination, panoramic radiography and dental histories | Overall  *Microdontia:*  30.6%  *Tooth agenesis:*  20.4%  *Mild enamel hypoplasia*:  5.1%  *Severe enamel hypoplasia*  7.1%  Stratified by age group at diagnosis  ***Microdontia***  < 3 years at dx: 57.8%  3-5 years at dx: 29.5%  > 5 years at dx: 11.4%  p < 0.001  ***Tooth agenesis***  < 3 years at dx: 39.1%  3-5 years at dx: 15.9%  > 5 years at dx: 9.1%  p < 0.001  *Mild enamel hypoplasia*  < 3 years at dx: 6.3%  3-5 years at dx: 4.5%  > 5 years at dx: 4.5%  p = 0.879  ***Severe enamel hypoplasia***  < 3 years at dx: 17.2%  3-5 years at dx: 6.8%  > 5 years at dx: 0%  p<0.001 |  |
| Quispe et al, 2019^7^ | Retrospective case-control study | Brazil | 97 CCS treated with chemotherapy and/or radiotherapy  111 healthy age- and sex-matched controls | Age at study:  CCS: 13.3 years  CG: age-matched  Age at diagnosis: 6.9 years | Panoramic radiographs | ***Microdontia***  CCS: 18.56%  CG: 0.90%  p < 0.001  ***Hypodontia***  CCS: 11.34%  CG: 3.60%  p = 0.026 |  |
| **Population-based, large studies with adult childhood cancer survivors** | | | | | | | |
| Kaste et al., 2009^8^ | Retrospective | USA & Canada | 8522 CCS diagnosed < 21 years who survived > 5 years  CG: 2831 siblings | Age at study: 44% 17-29 years, 56% ≥ 30 years  Age at dx: 6 years | Self-report | *Microdontia*  CCS: 9.2%  CG: 3.3%  *Hypodontia*  CCS: 8.2%  CG: 5.3%  *Enamel hypoplasia*  CCS: 11.7%  CG: 5.3%  *> 5 cavities*  CCS: 52%  CG: 51%  *Gingivitis*  CCS: 6.7%  CG: 5.7% | CCS compared to siblings  Models were adjusted for sex, race, education, household income, health insurance, and age at follow-up  ***Microdontia***  CCS compared to siblings: OR = 3.0, 95%CI 2.4-3.8  ***Hypodontia***  CCS compared to siblings: OR = 1.7, 95%CI 1.4-2.0  ***Enamel hypoplasia***  CCS compared to siblings: OR = 2.4, 95%CI 2.0-2.9  ***> 5 cavities***  CCS compared to siblings: OR = 1.2, 95%CI 1.1-1.3  ***Gingivitis***  CCS compared to siblings: OR = 1.2, 95%CI 1.0-1.5 |
| Patni et al., 2023^9^ | Retrospective | USA | 4865 CCS treated at St. Jude Children’s Research Hospital (SJLIFE Cohort)  CG: 591 community controls | Age at study: CCS: 31 years (23-39)  CG: 32 years (25-41)  Age at dx: 41% 0-5 years, 22% 5-10 years, 21% 10-15 years, 16% > 15 years | Self-report | *Microdontia*  CCS: 14%  CG: 2.1%  *Hypodontia*  CCS: 10%  CG: 4%  *Enamel hypoplasia*  CCS: 17%  CG: 4.6%  *> 5 cavities*  CCS: 49%  CG: 47%  *Gingivitis*  CCS: 10%  CG: 5.3% | CCS compared to siblings  Models were adjusted for sex, race, education, household income, health insurance, and age at follow-up  ***Microdontia***  CCS compared to siblings: OR = 7.89, 95%CI 4.64-14.9  ***Hypodontia***  CCS compared to siblings: OR = 2.75, 95%CI 1.83-4.33  ***Enamel hypoplasia***  CCS compared to siblings: OR = 4.24, 95%CI 2.90-6.49  *> 5 cavities*  CCS compared to siblings: OR = 1.11, 95%CI 0.93-1.33  ***Gingivitis***  CCS compared to siblings: OR = 2.04, 95%CI 1.43-3.03 |

*CCS = childhood cancer survivors; CG = Control group; DMFT = decayed missing filled teeth; dx = diagnosis*

### **Supplementary Figure 1.** Study population tree

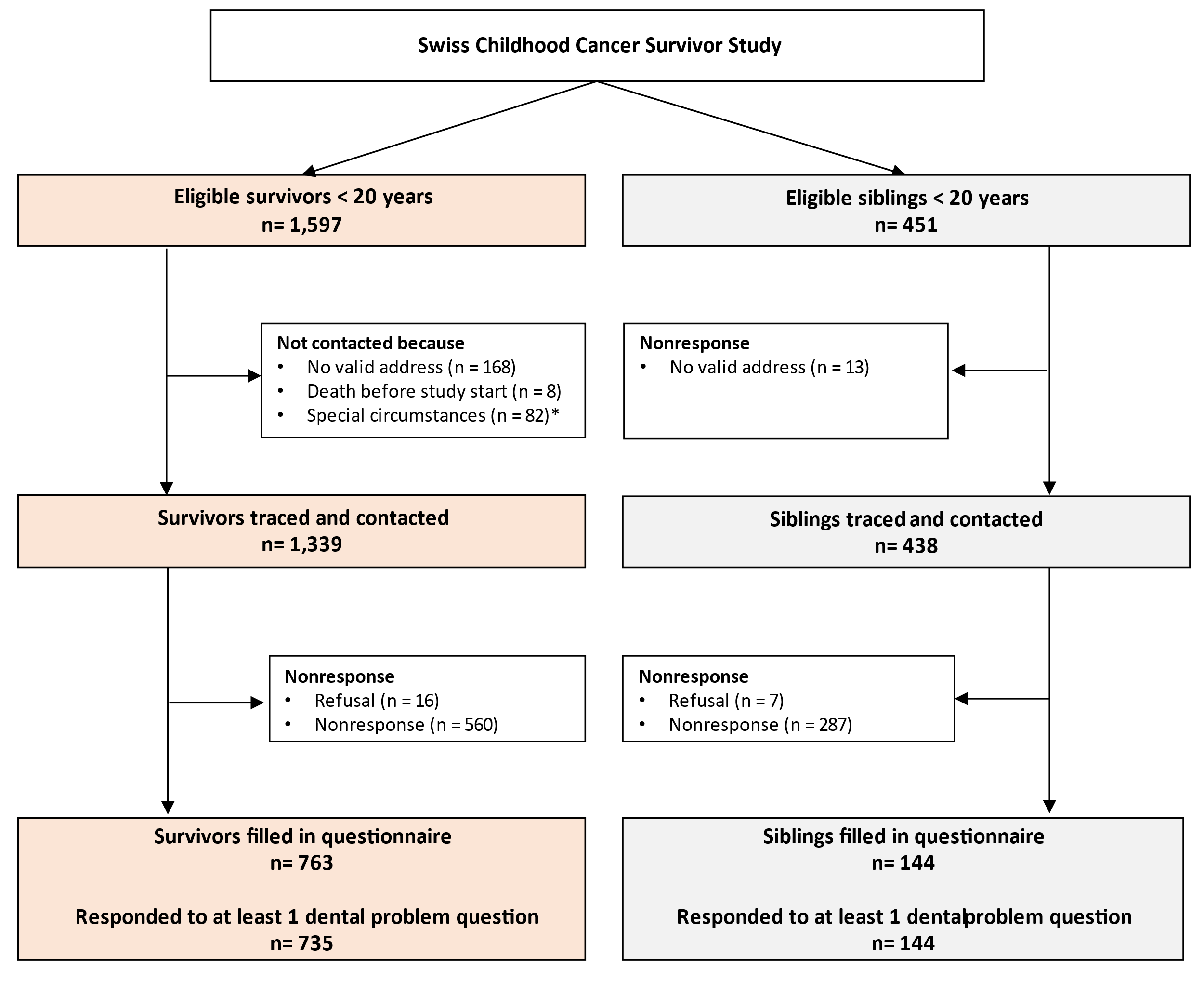

** Special circumstances include reasons that made it inappropriate to contact the childhood cancer survivor (e.g., relapse at the time of the study)*

**Supplementary Table 1.** Overview dental health questions, response options, and categorization

| **Dental health problem** | **Question / statement in the questionnaire** | **Response option** | **Coding for analysis** |
| --- | --- | --- | --- |
| Hypodontia | One/multiple teeth did not develop | No/yes | 0: no  1: yes |
| Microdontia | Underdeveloped/malformed teeth | No/yes | 0: no  1: yes |
| Enamel hypoplasia | Missing tooth enamel | No/yes | 0: no  1: yes |
| Cavities | Number of teeth that had to be fixed due to cavities | Participants reported the number of teeth in a free text field | 0: no (0 teeth)  1: yes (≥ 1 teeth) |
| Cavity-related tooth loss | Number of teeth lost due to cavities or gum problems | Participants reported the number of teeth in a free text field | 0: no (0 teeth)  1: yes (≥ 1 teeth) |
| Gum problems during cancer treatment^a^ | Gum problems during cancer treatment | Never / sometimes / often | 0: no (never)  1: yes (sometimes/often) |
| Gum problems after cancer treatment^a^ | Gum problems after cancer treatment | Never / sometimes / often | 0: no (never)  1: yes (sometimes/often) |

*^a^ only asked in childhood cancer survivor questionnaire, not in siblings*

### **Supplementary Table 2.** Prevalence of dental problems in childhood cancer survivors and siblings

|  | **Survivors** | | | |  | **Siblings** | | | |  |
| --- | --- | --- | --- | --- | --- | --- | --- | --- | --- | --- |
|  | *Total N^a^* | *N^b^* | *%* | *95%CI* |  | *Total N^a^* | *N^b^* | *%^c^* | *95%CI^c^* | *p^d^* |
| At least 1 dental problem | 735 | 329 | 45 | 41;48 |  | 144 | 66 | 47 | 38;57 | 0.609 |
| *Hypo- or microdontia^e^* | *731* | *103* | *14* | 12;17 |  | *144* | *14* | 9 | 4;13 | 0.089 |
| Hypodontia | 725 | 55 | 8 | 6;10 |  | 144 | 8 | 4 | 1;7 | 0.093 |
| Microdontia | 714 | 76 | 11 | 8;13 |  | 144 | 6 | 5 | 1;9 | **0.048** |
| Enamel hypoplasia | 706 | 55 | 8 | 6;10 |  | 144 | 5 | 4 | 0;7 | 0.094 |
| *Cavities or tooth loss^f^* | *724* | *266* | *37* | *33;40* |  | *144* | *54* | *41* | *31;50* | 0.439 |
| Cavities that had to be fixed | 642 | 249 | 39 | 35;43 |  | 138 | 53 | 42 | 33;52 | 0.491 |
| Tooth loss | 694 | 39 | 6 | 4;7 |  | 136 | 7 | 4 | 1;7 | 0.346 |
| Gum problems during treatment | 676 | 229 | 34 | 30;37 |  |  |  |  |  |  |
| Gum problems after treatment | 637 | 88 | 14 | 11;17 |  |  |  |  |  |  |

*^a^ Total N corresponding to the number of CCS / siblings responding to this question*

*^b^ Number of CCS / siblings reporting to have this problem*

*^c^ For siblings, we report the weighted percentages*

*^d^ Results of chi-square test based upon weighted proportions*

*^e^ Comprises participants reporting on hypodontia, microdontia or both*

*^f^ Comprises participants reporting on cavities that had to be fixed, tooth loss due to cavities/gum disease, or both*

### **Supplementary Table 3.** Overview dental problems in childhood cancer survivors stratified by age at diagnosis

|  | **CCS diagnosed < 5 years** | | | |  | **CCS diagnosed ≥ 5 years** | | | |  |
| --- | --- | --- | --- | --- | --- | --- | --- | --- | --- | --- |
|  | *Total N^a^* | *N^b^* | *%* | *95%CI* |  | *Total N^a^* | *N^b^* | *%* | *95%CI* | *p^c^* |
| At least one dental problem | 433 | 199 | 46 | 41;51 |  | 302 | 130 | 43 | 38;49 | 0.435 |
| *Hypo- or microdontia^d^* | 433 | 81 | 19 | 15;23 |  | 298 | 22 | 7 | 5;11 | **<0.001** |
| Hypodontia | 428 | 43 | 10 | 7;13 |  | 297 | 12 | 4 | 2;7 | **0.003** |
| Microdontia | 421 | 61 | 14 | 11;18 |  | 293 | 15 | 5 | 3;8 | **<0.001** |
| Enamel hypoplasia | 416 | 38 | 9 | 6;12 |  | 290 | 17 | 6 | 4;9 | 0.110 |
| *Cavities or tooth loss^e^* | 428 | 151 | *35* | *31;40* |  | 296 | 115 | *39* | *33;44* | 0.327 |
| Cavities that had to be fixed | 382 | 138 | 36 | 31;41 |  | 260 | 111 | 43 | 37;49 | 0.094 |
| Tooth loss | 411 | 27 | 7 | 5;9 |  | 283 | 12 | 4 | 2;7 | 0.190 |
| Gum problems during treatment | 397 | 119 | 30 | 26;35 |  | 279 | 110 | 39 | 34;45 | **0.011** |
| Gum problems after treatment | 371 | 44 | 12 | 9;16 |  | 266 | 44 | 17 | 13;22 | 0.091 |

*^a^ Total N corresponding to the number of CCS responding to this question*

*^b^ Number of CCS reporting to have this problem*

*^c^ Results of chi-square test*

*^d^ Comprises participants reporting on hypodontia, microdontia or both*

*^e^ Comprises participants reporting on cavities that had to be fixed, tooth loss due to cavities/gum disease, or both*

### **Supplementary Table 4.** Overview dental problems in childhood cancer survivor and siblings

|  | **Survivors**  **(N = 735)** | |  | **Siblings**  **(N = 144)** | |
| --- | --- | --- | --- | --- | --- |
| **Default questionnaire categories** | **N** | **%** |  | **N** | **%^b^** |
| **Hypodontia**  *One/multiple teeth did not develop* |  |  |  |  |  |
| No | 670 | 91% |  | 136 | 94% |
| Yes | 55 | 8% |  | 8 | 6% |
| *Missing* | *10* |  |  | *0* |  |
| **Microdontia**  *Underdeveloped/malformed teeth* |  |  |  |  |  |
| No | 638 | 87% |  | 138 | 96% |
| Yes | 76 | 10% |  | 6 | 4% |
| *Missing* | *21* |  |  | *0* |  |
| **Enamel hypoplasia** |  |  |  |  |  |
| *Too little or missing tooth enamel* |  |  |  |  |  |
| No | 651 | 89% |  | 139 | 97% |
| Yes | 54 | 7% |  | 5 | 4% |
| *Missing* | *30* |  |  | *0* |  |
| **Cavities that had to be fixed**  *Holes that had to be fixed due to cavities* |  |  |  |  |  |
| No | 393 | 54% |  | 85 | 59% |
| Yes | 249 | 34% |  | 53 | 37% |
| *Missing* | *93* |  |  | *6* |  |
| **Tooth lost due to cavities or gum disease** |  |  |  |  |  |
| *Tooth lost due to cavities or gum disease; tooth fell out or had to be pulled* |  |  |  |  |  |
| No | 655 | 89% |  | 129 | 90% |
| Yes | 39 | 5% |  | 7 | 5% |
| *Missing* | *41* |  |  | *8* |  |
| **Gum problems during treatment^a^**  *Gum problems during cancer treatment* |  |  |  |  |  |
| Never | 447 | 61% |  |  |  |
| Sometimes | 148 | 20% |  |  |  |
| Often | 81 | 11% |  |  |  |
| *Missing* | *59* |  |  |  |  |
| **Gum problems after treatment^a^**  *Gum problems after cancer treatment* |  |  |  |  |  |
| Never | 549 | 75% |  |  |  |
| Sometimes | 72 | 10% |  |  |  |
| Often | 16 | 2% |  |  |  |
| *Missing* | *98* |  |  |  |  |
| **Coded categories based upon qualitative responses from other tooth problems** | | | | |  |
| Orthodontic correction or jaw misalignment | 55 |  |  | 15 |  |
| Abnormal root development | 6 |  |  | 1 |  |
| First teeth stayed longer than normal | 3 |  |  | 0 |  |
| Colored teeth (yellow/brown) | 6 |  |  | 0 |  |
| Additional tooth / teeth | 1 |  |  | 1 |  |
| Tooth/teeth pulled (reason unspecified or other than caries) | 4 |  |  | 0 |  |
| Mouth cystitis or ulcerus | 3 |  |  | 0 |  |
| Aphthae (Speck, flake or blister in the mouth area) | 3 |  |  | 2 |  |

*^a^ Only asked in childhood cancer survivor questionnaires*

^b^ *Proportions without survey weights*

### **Supplementary Table 5.** Problems with health insurance coverage for dental problems of CCS who reported at least one dental problem (N = 329).

| **Dental problem** | **N** | **%** |
| --- | --- | --- |
| **Any dental problem (N = 329)** |  |  |
| No problems with health insurance coverage | 245 | 80% |
| Problem with health insurance coverage | 60 | 20% |
| *Missing* | *24* |  |
| **Hypodontia (N = 55)** |  |  |
| No problems with health insurance coverage | 40 | 80% |
| Problem with health insurance coverage | 10 | 20% |
| *Missing* | *5* |  |
| **Microdontia (N = 76)** |  |  |
| No problems with health insurance coverage | 57 | 73% |
| Problem with health insurance coverage | 14 | 27% |
| *Missing* | *5* |  |
| **Enamel hypoplasia (N = 55)** |  |  |
| No problems with health insurance coverage | 35 | 79% |
| Problem with health insurance coverage | 13 | 21% |
| *Missing* | *7* |  |
| **Cavities that had to be fixed (N = 249)** |  |  |
| No problems with health insurance coverage | 188 | 79% |
| Problem with health insurance coverage | 49 | 21% |
| *Missing* | *12* |  |
| **Tooth lost due to cavities or gum disease (N = 39)** |  |  |
| No problems with health insurance coverage | 23 | 62% |
| Problem with health insurance coverage | 14 | 38% |
| *Missing* | *2* |  |
| **Gum problems during treatment (N = 229)** |  |  |
| No problems with health insurance coverage | 187 | 87% |
| Problem with health insurance coverage | 29 | 13% |
| *Missing* | *13* |  |
| **Gum problems after treatment (N = 88)** |  |  |
| No problems with health insurance coverage | 66 | 79% |
| Problem with health insurance coverage | 18 | 21% |
| *Missing* | *4* |  |

*The N corresponds to the number of survivors reporting to have this problem.*

### **Supplementary Table 6.** Sensitivity analysis: Dental problems in childhood cancer survivors compared to siblings with missing data imputed (N = 907)

|  | **Hypo- or microdontia** | | | | **Enamel hypoplasia** | | | | **Fixed cavities or tooth loss** | | | |
| --- | --- | --- | --- | --- | --- | --- | --- | --- | --- | --- | --- | --- |
|  | *OR* | *SE* | *95%CI* | *p* | *OR* | *SE* | *95%CI* | *p* | *OR* | *SE* | *95%CI* | *p* |
| CCS (ref. sibling) | 1.73 | 0.33 | 0.91;3.29 | 0.093 | 2.31 | 0.50 | 0.86;6.17 | 0.095 | 0.85 | 0.21 | 0.56;1.28 | 0.442 |
| Age at study | 0.94 | 0.04 | 0.87;1.01 | 0.070 | 0.95 | 0.04 | 0.88;1.03 | 0.239 | 0.98 | 0.03 | 0.92;1.04 | 0.544 |
| Sex (ref. girls) | 0.89 | 0.27 | 0.52;1.52 | 0.672 | 0.91 | 0.36 | 0.45;1.85 | 0.797 | 1.05 | 0.21 | 0.70;1.58 | 0.818 |

### **Supplementary Table 7.** Sensitivity analysis: Cancer-related risk factors of dental problems in childhood cancer survivors in multivariable logistic regression with missing data imputed (N = 763)

|  | **Hypo- or microdontia** | | | | **Enamel hypoplasia** | | | | **Fixed cavities or tooth loss** | | | |
| --- | --- | --- | --- | --- | --- | --- | --- | --- | --- | --- | --- | --- |
|  | *OR* | *SE* | *95%CI* | *p* | *OR* | *SE* | *95%CI* | *p* | *OR* | *SE* | *95%CI* | *p* |
| Cranial radiation (ref. no) | 1.13 | 0.39 | 0.53;2.40 | 0.759 | 0.93 | 0.51 | 0.34;2.54 | 0.883 | 0.86 | 0.29 | 0.49;1.52 | 0.637 |
| Chemotherapy (ref. no) | 2.21 | 0.36 | 1.10;4.45 | **0.027** | 2.83 | 0.52 | 1.02;7.90 | **0.046** | 1.48 | 0.21 | 0.98;2.23 | **0.049** |
| HSCT (ref. no) | 2.51 | 0.32 | 1.35;4.68 | **0.004** | 1.87 | 0.39 | 0.87;4.03 | 0.110 | 2.09 | 0.27 | 1.23;3.56 | **0.006** |
| Age at diagnosis | 0.84 | 0.04 | 0.78;0.91 | **<0.001** | 0.91 | 0.05 | 0.83;1.00 | 0.054 | 1.04 | 0.02 | 0.99;1.09 | 0.094 |
| Time since diagnosis | 1.11 | 0.05 | 1.00;1.23 | 0.051 | 1.03 | 0.07 | 0.89;1.19 | 0.732 | 1.12 | 0.04 | 1.03;1.22 | **0.005** |
| Relapse (ref. no) | 1.00 | 0.35 | 0.50;2.00 | 0.992 | 2.17 | 0.38 | 1.03;4.58 | **0.043** | 0.62 | 0.28 | 0.36;1.08 | 0.082 |
|  | **Gum problems during treatment** | | | | **Gum problems after treatment** | | | |  | | | |
|  | *OR* | *SE* | *95%CI* | *p* | *OR* | *SE* | *95%CI* | *p* |  |  |  |  |
| Cranial radiation (ref. no) | 1.57 | 0.29 | 0.89;2.77 | 0.117 | 1.45 | 0.34 | 0.75;2.81 | 0.274 |  |  |  |  |
| Chemotherapy (ref. no) | 21.88 | 0.49 | 8.29;57.71 | **<0.001** | 4.75 | 0.46 | 1.92;11.77 | **0.001** |  |  |  |  |
| HSCT (ref. no) | 2.10 | 0.28 | 1.21;3.67 | **0.009** | 1.70 | 0.33 | 0.89;3.24 | 0.105 |  |  |  |  |
| Age at diagnosis | 1.11 | 0.03 | 1.06;1.17 | **<0.001** | 1.09 | 0.03 | 1.02;1.16 | **0.013** |  |  |  |  |
| Time since diagnosis | 1.03 | 0.05 | 0.93;1.13 | 0.609 | 1.15 | 0.06 | 1.02;1.28 | **0.018** |  |  |  |  |
| Relapse (ref. no) | 0.99 | 0.29 | 0.56;1.73 | 0.964 | 1.44 | 0.35 | 0.72;2.89 | 0.303 |  |  |  |  |

**References**

1.	Busenhart DM, Erb J, Rigakos G, Eliades T, Papageorgiou SN. Adverse effects of chemotherapy on the teeth and surrounding tissues of children with cancer: A systematic review with meta-analysis. *Oral Oncol* 2018; **83**: 64-72.

2.	Pombo Lopes J, Rodrigues I, Machado V, Botelho J, Bandeira Lopes L. Chemotherapy and Radiotherapy Long-Term Adverse Effects on Oral Health of Childhood Cancer Survivors: A Systematic Review and Meta-Analysis. *Cancers* 2023; **16**(1): 110.

3.	Seremidi K, Kloukos D, Polychronopoulou A, Kattamis A, Kavvadia K. Late effects of chemo and radiation treatment on dental structures of childhood cancer survivors. A systematic review and meta‐analysis. *Head & Neck* 2019; **41**(9): 3422-33.

4.	Defabianis P, Bocca N, Romano F. Prevalence and association of dental anomalies and tooth decay in Italian childhood cancer survivors. *J Clin Pediatr Dent* 2023.

5.	Halperson E, Matalon V, Goldstein G, et al. The prevalence of dental developmental anomalies among childhood cancer survivors according to types of anticancer treatment. *Sci Rep* 2022; **12**(1).

6.	Kang C-M, Hahn SM, Kim HS, et al. Clinical Risk Factors Influencing Dental Developmental Disturbances in Childhood Cancer Survivors. *Cancer Res Treat* 2018; **50**(3): 926-35.

7.	Quispe RA, Rodrigues ACC, Buaes AMG, Capelozza ALA, Rubira CMF, Santos PSdS. A case-control study of dental abnormalities and dental maturity in childhood cancer survivors. *Oral Surgery, Oral Medicine, Oral Pathology and Oral Radiology* 2019; **128**(5): 498-507.e3.

8.	Kaste SC, Goodman P, Leisenring W, et al. Impact of radiation and chemotherapy on risk of dental abnormalities. *Cancer* 2009; **115**(24): 5817-27.

9.	Patni T, Lee C-T, Li Y, et al. Factors for poor oral health in long-term childhood cancer survivors. *BMC Oral Health* 2023; **23**(1): 73.
